# Epidemiology of Door Crush Injuries- A retrospective cohort study of South Indian Population in a Tertiary care center

**DOI:** 10.1101/2022.05.13.22274648

**Authors:** Ashwin Krishnamoorthy, Maithreyi Sethu, Singaravelu Viswanathan, Srinivasan Rajappa, Sathishkumar Jayaram

## Abstract

More than half of the fingertip injuries in children are due to door jamming injuries.^1)2)^ There have been several studies on door crush injuries (DCI) but they pertain to either the paediatric age group or form a part of study of fingertip injuries in a large population. This article caters solely to studying the epidemiology, mechanism of injury, associated risk factors and suggests few simple techniques to avoid DCI.

**Materials:** Comparative analysis of the epidemiological data of all the patients with door crush injuries who presented to the Emergencies and the Out Patient Department in the Tertiary care centre was obtained from the MRD. This is a retrospective cohort study between January 2021 to December 2021. Patients with serious concomitant injuries, machine crush injury, heavy falling objects, window crush injury were excluded from the study.

**Results:** Of the 34 patients, 27 were male and 7 females. In 33 patients DCI was in the hinge side while only 1 had lock-side, entrance door being the commonest. 35% had first-aid done in a local nursing home before arriving at the hospital. 4 patients left against medical advice, 4 were conservatively treated and 2 had double finger injury. DCI was most common in preschool children. Right side and middle finger were most susceptible. 25% of the injuries happened on Mondays. Of the 36 fingers injured, 69% had pulp, 58% had nailbed and 22% had bony involvement. Primary suturing, local flap cover and K-wiring were the main modalities of treatment. Complications included altered sensation, nail deformity and contracture.

**Conclusion:** Door crush injury is a major contributor for finger crush injuries in both children and adults. Awareness among parents, the use of safety appliances to prevent accidental door closing, counselling by doctors and nurses greatly help to bring down the number of door crush injuries.

## Background

Doors have been an integral part of human evolution from the time of Egyptians – about 4000 years ago. Sliding and double doors have existed in Roman temples as early as 79 AD. Doors continue to form a vital part of our lives providing us privacy, security and aesthetic benefit. Associated with the advent of doors was the rise in door-related injuries. They came to be known popularly as door crush injuries (DCI). DCI commonly cause fingertip injuries in children as well as adults.

There are two main varieties of DCI – lock side and the hinge side door crush injuries. It has been found that younger children (<10 years) tend to crush their fingers more on the hinge side (53%) and older children (>10 years) on the lock side (55%) of the door.^3) 1)^

Hand injuries account for nearly 10% of all cases in emergency departments (ED).^4)^ In the paediatric population, it is found that hand injuries accounted for 1.8-2%^5) 6)^ of attendance in the children’s emergency department and out of these, 21-46% were fingertip injuries.^6) 7)^Nearly half or more of the fingertip injuries in children were due to door jamming injuries. ^1)2)^

Approximately 4.8 million emergency visits in the USA are attributed to fingertip injuries. India does not have similar statistics,^8)^ yet these voluminous figures portray an idea of the magnitude of DCI in all parts of the world.

There have been several studies on DCI^5, 8–10)^ but they pertain to either the paediatric age group or form a part of study of fingertip injuries in a large population. This article caters solely to studying the epidemiology, possible mechanism of injury, associated risk factors and suggests few simple techniques to avoid door crush injuries.

### Patients and Methods

The data of all the patients with door crush injuries who presented to the Emergency and Out Patient Departments in a Tertiary care centre in South India was obtained from the Medical Record Department. This is a retrospective study spanning over one year from January 2021 to December 2021.

#### Objectives

To study the epidemiology of door crush injuries in a tertiary care centre in South India.

#### Inclusion Criteria

Patients of all age groups with door crush injuries within 2 days of the trauma.

#### Exclusion Criteria

Patients with concomitant serious injuries, other aetiologies of crush such as machine crush injury of heavy falling objects and cut injuries, window crush injury were excluded from the study.

Patients with chronic wounds and those treated for the door crush injury in other hospitals were also excluded.

All the surgeries were performed by highly experienced surgeons, from the departments of Hand Surgery and Plastic Surgery. The patients were followed for a period of 3 months minimum. Recall bias was avoided by documenting all the findings during the admission and the OPD visits in the follow-up period using tabulated proformas. Loss of follow-up was minimised by telemedicine using audio-visual phone calls. The study was approved by the Institutional Ethical Committee. All the data was collected and analysed using ratios, graphs and charts to calculate the statistics of the epidemiology.

## Results

There were 34 patients in total and all of them presented in the Emergency Department.

- Most common age group for door crush injury was between 0-5 years which constituted 35% of the case load. There was also another peak in the 21-25 years age group making up 14.7% of the total population of DCI.
- It was found that (Fig 1) in the first three years of age the incidence of DCI was maximum.
- The Male: Female sex ratio was found to be 27:7.
- Two patients had DCI involving two fingers. In both scenarios middle and ring fingers were injured, the rest had only single finger injury.
- Ratio of percentage of finger involvement
- DCI were more common on the right side, 21 right vs 13 left.
- Mode of injury: Hinge side injuries were predominant as compared to the lock side with 33 on the side of the Hinge and only 1 on the lock-side.
- Car door vs house door (Table 1): 11.8% of the injuries were due to vehicle door crush while a majority 76% was due to door crush injury in their residence. Of these, 44% were entrance doors and 21%-bedroom doors.
- Place of injury: Home was the most common site at which the injury occurred. This is most probably due to Covid when many people were working from home and there was no school for most of the study period.
- Time during the day: Most common times of injury were 3 pm and 9 pm when around 12% if the injuries occurred.
- There was a peak of DCI on Mondays, constituting around 1/4 of the cases (23.5%) (Table2).
- The most common first aid given was dressing in local nursing homes for achieving haemostasis (35%) followed by applying ice packs (14%) (table 3).
- 69% of the patients had injured their pulp which needed at least one suture, 58% had nail bed injury while only 22% had sustained bony injury (Table 4).
- There were 3 patients who had fracture proximal to the fingertip. First with a distal and middle phalanx fracture of the injured ring finger, second with a terminal phalanx base fracture of the little finger and the third with a middle phalanx fracture on the injured little finger.
- 27.78% of the DCI were amputations or sub-total amputations (Table 5).
- 11.8% of the patients were treated conservatively and another 11.8% refused treatment and left against medical advice.
- Pulp and Nail Bed Suturing were the most common surgical treatment given followed by Flap cover and shortening of the finger (Table 6). Replant was not a viable requirement or option for any of the patients.
- Two patients had altered sensation-one hyperaesthesia (was dissatisfied with the treatment) and one with reduced sensation at the end of 3 months after the treatment, one patient required secondary suturing and one patient had nail plate deformity (Hook Nail). One patient who developed flexion contracture of distal interphalangeal joint was dissatisfied with the loss of function of the joint.

**Figure 1:**
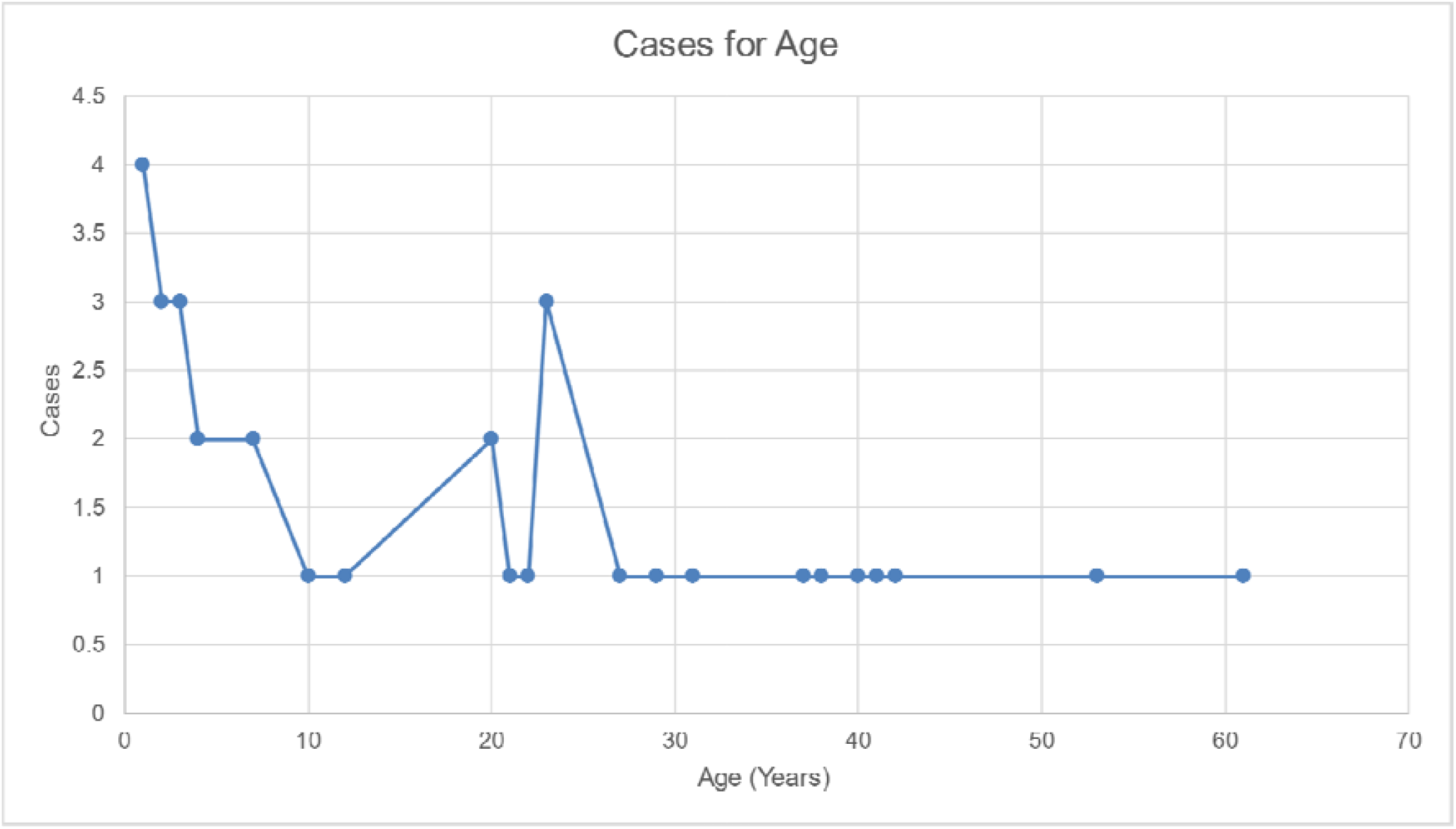
No. of cases of DCI vs Age

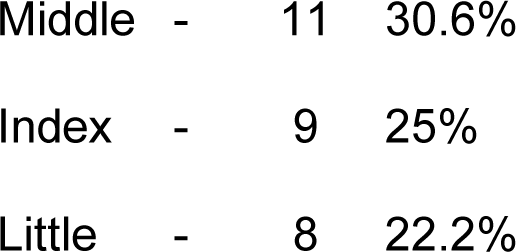
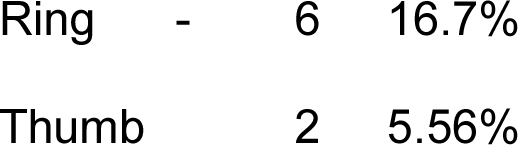

**Table 1:** Doors involved in door crush injuries

| DOOR INVOLVED<br>IN THE INJURY | NO. OF<br>CASES | PERCENTAGE OF CASES |
| --- | --- | --- |
| Bathroom | 4 | 11.76 |
| Entrance | 15 | 44.12 |
| Bedroom | 7 | 20.59 |
| Car Door | 3 | 8.82 |
| Bus | 1 | 2.94 |
| Gate | 4 | 11.76 |
| Total | 34 | 100 |

**Table 2:** Door crush injury vs day of the week

| DAY OF WEEK | NO. OF CASES | PERCENTAGE |
| --- | --- | --- |
| Monday | 8 | 23.5 |
| Tuesday | 2 | 5.9 |
| Wednesday | 5 | 14.7 |
| Thursday | 4 | 11.8 |
| Friday | 6 | 17.6 |
| Saturday | 3 | 8.8 |
| Sunday | 6 | 17.6 |
| Total | 34 | 100 |

**Table 3:**
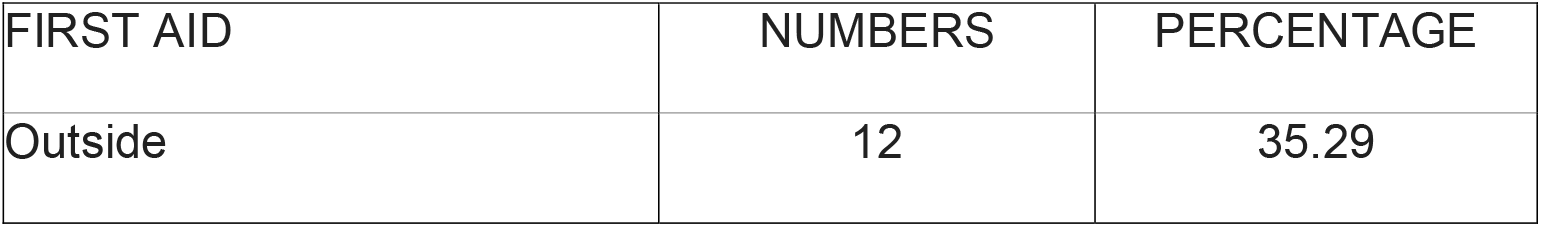
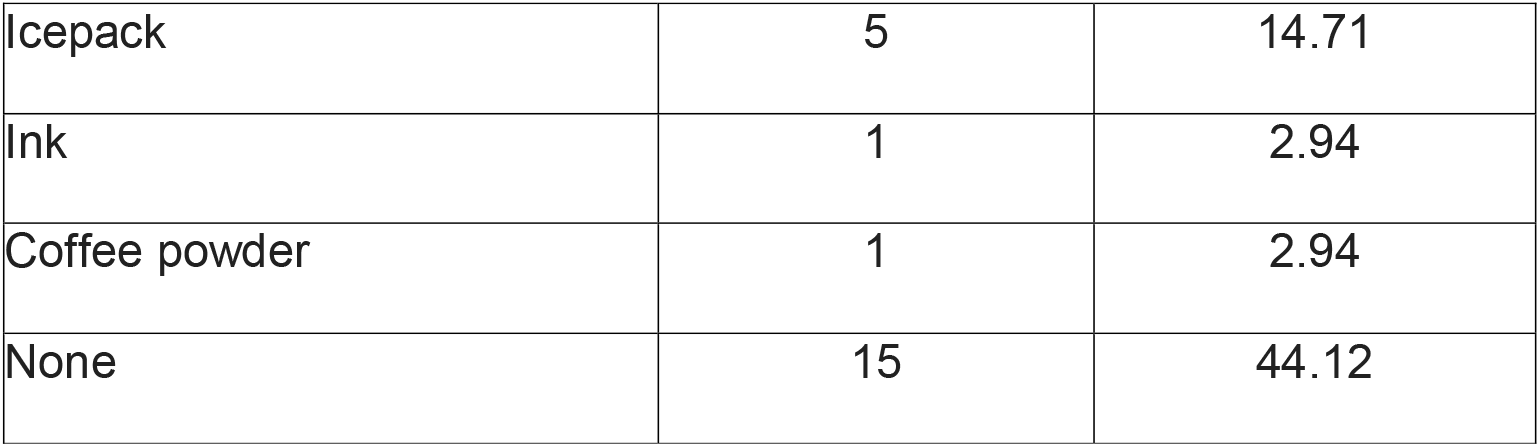
First Aid

| FIRST AID | NUMBERS | PERCENTAGE |
| --- | --- | --- |
| Outside | 12 | 35.29 |
| Icepack | 5 | 14.71 |
| Ink | 1 | 2.94 |
| Coffee powder | 1 | 2.94 |
| None | 15 | 44.12 |

**Table 4:** Injured components of the finger

| INJURED COMPONENT | NO. OF CASES | PERCENTAGE |
| --- | --- | --- |
| PULP (inclusive of all pulps injuries) | 25 | 69.44 |
| NAIL BED(inclusive of all nailbed injuries) | 21 | 58.33 |
| BONE(Inclusive of all bony injuries) | 8 | 22.22 |
| PULP +NAILBED +BONE | 6 | 16.67 |
| PULP +NAILBED | 11 | 30.56 |
| PULP+BONE | 1 | 2.78 |
| NAILBED +BONE | 1 | 2.78 |
| DORSAL SOFT TISSUE INJURY/LOSS | 1 | 2.78 |

**Table 5:** Amputation levels in door crush injuries

| ALLEN TYPE | CASES | PERCENTAGE IN TOTAL CASES |
| --- | --- | --- |
| ALLEN I | 1 | 2.78 |
| ALLEN II | 4 | 11.11 |
| ALLEN III | 3 | 8.33 |
| ALLEN IV | 2 | 5.56 |

**Table 6:** Surgical treatment for door crush injuries

| TREATMENT | NO OF CASES | PERCENTAGE OF CASES |
| --- | --- | --- |
| Pulp Suturing | 23 | 67.65 |
| Nail Bed Suturing | 20 | 58.82 |
| Flap | 6 | 11.65 |
| Shortening closure | 2 | 5.88 |
| Nail bed grafting | 1 | 2.94 |
| Composite graft | 1 | 2.94 |
| K wire | 1 | 2.94 |
| Skin graft | 1 | 2.94 |

**Table 7:** Complications of door crush injury

| COMPLICATIONS | NUMBER | PERCENTAGE | DISSATISFIED(Nos) |
| --- | --- | --- | --- |
| Altered Sensation | 2 | 7.14 | 1 |
| Secondary suturing | 1 | 3.57 | - |
| Nail Deformity | 1 | 3.57 | - |
| Contracture | 1 | 3.57 | 1 |

## Discussion

The incidence of door crush injury was found to be prevalent among the paediatric age group 1-4 yrs. Several studies have indicated that maximum door crush injuries and fingertip injuries occurred at 5 – 6 years age. ^5)11)10)^ In the present study, a smaller, second peak was seen in the 20-25 years age group, where the individuals seemed to be in a hurry when the accident happened.

Predominant DCIs were sustained by males when compared to females in both the paediatric and adult population. This has been established in several other studies too where males constituted around 60-70% of the paediatric patients.^4) 6) 2) 9)^ Paediatric age group of less than 5 years and male sex seem to a non-modifiable risk factor for door crush injuries pointing towards the need for a better-quality supervision by the parents.

Middle finger was the commonest finger to be injured. This is concurrent with other studies on paediatric door crush injuries and is being attributed to the length of the finger. ^8), 11)^

On comparing the sides, right sided injuries seem to be more common that the left (23 vs 13) in all age groups.

Except one patient, all sustained injury at the side of the hinge and in most of the paediatric cases the person who closed the door was not aware of the child standing on the other side with the finger at the hinge of the door. There were also two incidences of the wind suddenly closing in on the door when the patient was placing his finger on the hinge. Other studies have also reported that hinge side is commoner for DCI as the child is not under direct vision of the person closing the door.^7)^

The comparison of DCI during vacation or at schools and offices could not be computed as we attribute most of the accidents happening in residences during the times of Covid lockdowns when children and most adults were home bound for most of the period.

Nearly 1/4^th^ of the injuries happened on Monday, in the beginning of the week when the anxiety and stress levels are generally higher after the relaxing weekend.

A complex injury is defined as either an injury to more than one of the anatomical components of the hand (bone, flexor/extensor tendon, joint, nerve and arteries) or total/subtotal amputations through the middle or proximal phalanges.^2)^ In this study, 70% of the patients had pulp injuries and 59% had nail bed injuries. This statistic is similar to that of Claudet et al where nail plate was damaged in 60% of digital lesions.^11)^

Around 29% of the injuries were associated with fracture of the terminal phalanx. 5.9% of the patients also had fracture of the proximal or middle phalanx of the finger. Studies show that the misdiagnosis rate of hand fractures is 8% with the leading cause of misdiagnosis being misinterpretation of epiphyses as fractures followed by missing multiple fractures.^12)^ It is important to remember to clinically examine the entire hand as the chances of missing out on a fracture is very high in a DCI due to these reasons.

25% of the patients had sustained amputations out of which 33.3% was Allen 2 and 33.3% was Allen 3. Studies show that finger amputations accounted for up to 91.6% of all paediatric traumatic amputations.^6)^ And amputations contribute to 0.84% of hand injuries.^4)^

Suboptimal management of these injuries can result in persistent pain, abnormal sensation, finger shortening, nail deformity, joint stiffness, and reduced grip strength.^8)^ Only one of the 34 patients in our series developed the complication of flexion contracture of the distal interphalangeal joint and had restriction of movement of the joint. 20.5% (7) of the patient noticed shortening of the finger, 5.8% (2) had altered sensation in the fingertip, 2.9% (1) patient developed a nail deformity and overall, two patients were dissatisfied with the result. Limitation of the study is that around six months of study period was during either partial or complete lockdown for COVID.

### Prevention and Implication

There are no regulatory bodies for advocating safety measures to prevent DCI even in developed countries. But there a few commercially available items which might help reduce their incidence such as the rubber stopper in door at lock side, triangle shaped plastic stopper at bottom of door preventing closure, Australian plastic door guard at hinge side, Danish “pinch free” door. ^3)^ Along with the safety door closure systems coupled with improved supervision, child-safety counselling by doctors and nurses should become a routine.^4)^

## Conclusion

Door crush injury is a major contributor for finger crush injuries and contributes a large percentage of upper limb amputations. Preschool is the most vulnerable age group though it is also seen in adults. Majority of patients are males. The most common mechanism of injury is accidentally closing the door without the awareness of a person on the other side with the finger placed in the hinge. Middle finger is the most commonly injured finger. Nearly a fourth of the DCIs are amputations, some not very noticeable and some more serious leading to social stigma. As with all problems, prevention is better than cure. Awareness among parents and caregivers, routine counselling by doctors (paediatricians) and nurses, use of protective and preventive gadgets to prevent sudden accidental closure of doors are found to be useful and can bring down the incidence of DCIs.

## Data Availability

All data produced in the present work are contained in the manuscript

## Acknowledgements

None

## Funding

This research received no specific grant from any funding agency in the public, commercial or not-for-profit sectors.

## Conflict of Interest

None

## Notes

### Competing Interest Statement

The authors have declared no competing interest.

### Funding Statement

This study did not receive any funding

### Author Declarations

Sri Ramachandra Institute of Higher Education and Research, India - Institutional Ethics Committee

